# Molecular Epidemiology of Diarrhoeagenic *Escherichia coli* in Africa: A Systematic Review and Meta-Analysis

**DOI:** 10.1101/2023.10.11.23296874

**Authors:** John Bosco Kalule, Linda A. Bester, Daniel L. Banda, Firehiwot Abera Derra, Francis Chikuse, Sofonias K. Tessema, Africa PGI Foodborne Diseases Genomic Surveillance Focus Group, Ebenezer Foster-Nyarko

## Abstract

**Introduction:** Diarrhoeagenic *Escherichia coli* (DEC) persistently challenges public health in Africa, contributing substantially to the diarrhoeal disease burden. This systematic review and meta-analysis illuminate the distribution and antimicrobial resistance (AMR) patterns of DEC pathotypes across the continent.

**Methods:** The review selectively focused on studies reporting prevalence and/or AMR of human-derived DEC pathotypes from African nations, excluding data from extra-intestinal, animal, and environmental sources, and studies focused on drug and mechanism experiments. Employing a robust search strategy, pertinent studies were retrieved from SCOPUS, PubMed, and EBSCOhost, processed with Covidence, and screened in alignment with PRISMA guidelines.

**Results:** The reviewed studies were predominantly hospital-based (80%) and paediatric-focused (91%), with a meagre 4.4% documenting DEC outbreaks. Seven DEC pathotypes were discerned, with Enteroaggregative *E. coli* (EAEC) being notably prevalent (43%, 95% CI: 30% – 55%) and Enteroinvasive *E. coli* (EIEC) least prevalent (24%, 95%CI: 17% – 32%). Identified non-susceptibilities were noted against essential antibiotics, including ciprofloxacin, ceftriaxone, and ampicillin, while instances of carbapenem and Extended-Spectrum Beta-Lactamase (ESBL) resistance were scarce.

**Conclusion:** Despite sporadic data on DEC prevalence and AMR in Africa, particularly in community settings, a palpable gap remains in real-time outbreak surveillance and comprehensive data documentation. Augmenting surveillance and embracing advancements in molecular/genomic characterisation techniques are crucial to precisely discerning the actual impact and resistance continuum of DEC in Africa.

## Background

Diarrhoea is a significant public health concern in sub-Saharan Africa—with a high incidence due to factors like limited access to clean water and sanitation—leading to millions of cases annually—and is exacerbated by limited healthcare access, particularly among young children, HIV-positive individuals, and visitors from abroad [1–3]. Diarrhoea manifests as a symptom originating from infections induced by various organisms, including bacteria, viruses, and parasites, predominantly propagated through water contaminated with faeces. In low-income nations, Rotavirus and *Escherichia coli* are two predominant causative agents of moderate-to-severe diarrhoea, along with other pathogens like *Cryptosporidium* and *Shigella* [4, 5].

Despite recent studies in Africa revealing the problematic emergence of antimicrobial resistance for common causes of diarrhoea such as diarrhoeagenic *E. coli*, the full scope, distribution, molecular epidemiology, and antimicrobial resistance of diarrhoeagenic bacterial pathogens in the continent remain poorly understood, mainly because many cases go undetected, unreported, and, consequently, untreated. A recent PulseNet International survey emphasised the absence of Whole Genome Sequencing (WGS) in foodborne surveillance outside the United States, Canada, and Europe, spotlighting significant disparities in resources and expertise across regions [6].

In response to this pressing need, the Africa Pathogen Genomics Initiative (PGI) of the Africa Centres for Disease Control and Prevention (Africa CDC) established a technical focus group of experts on Foodborne Diseases (FBD) in April 2022. A significant area of concern is *E. coli*, a member of the *Enterobacteriaceae* family, which the World Health Organisation (WHO) has identified as one of twelve bacterial families that significantly threaten human health due to escalating antibiotic resistance [7]. The most vulnerable, such as young children, older adults, and those with compromised immunity or malnutrition, are at heightened risk. Key transmission factors include unhygienic practices, limited sanitation, and exposure to contaminated water sources for consumption and irrigation. The latter has been pinpointed as a significant factor in transmitting genes related to antibiotic resistance and increased pathogenicity [8].

While current research primarily analyses *E. coli* samples from diarrhoeic patients, there is a significant gap in our understanding of its prevalence in the broader community setting [1, 9]. The Global Enteric Multicenter Study (GEMS) provided insights into the genomic diversity of *E. coli*. Among others, their findings suggest the potential for certain strains to carry or acquire virulence genes typically associated with *E. coli* diarrhoeagenic pathotypes [10–12]. Beyond these insights, a fragmented understanding of *E. coli* pathotypes and their contribution to diarrhoeal diseases across the continent remains. Consequently, we lack a cohesive picture of this pivotal pathogen’s epidemiology and associated antibiotic resistance in African settings.

Contrasting with developed regions such as the USA and Europe, which have robust *E. coli* surveillance systems [13–18], Africa contends with significant systemic challenges. The value of well-established FBD surveillance systems was exemplified by the United Kingdom’s swift containment of a Shiga toxin-producing *Escherichia coli* (STEC) outbreak within five weeks using WGS [19]. As plans unfold for an African genomic FBD surveillance platform, understanding the prevalence, burden, and diversity of diarrhoeagenic *E. coli* from Africa becomes imperative. Addressing this gap is crucial, as it informs where to allocate resources and infrastructure.

Consequently, this systematic review examines the existing literature on diarrhoeagenic *E. coli* obtained from human stool samples of diarrhoeic cases in African healthcare settings and communities. Our objective is to elucidate the status of the main diarrhoeagenic *E. coli* pathotypes, viz. enteropathogenic *E. coli* (EPEC), Shiga toxin-producing *E. coli* (STEC), enteroaggregative *E. coli* (EAEC), enterotoxigenic *E. coli* (ETEC), enteroinvasive *E. coli* (EIEC), and diffusely adherent *E. coli* (DAEC) and their antibiotic resistance profiles, which directly challenge primary therapeutic measures. By aggregating data until December 2022, this review sets the foundation for developing a comprehensive pan-African surveillance system that integrates WGS insights.

## Methods

This systematic review utilised the Covidence (Veritas Health Innovation Ltd) data management system and adhered to the Preferred Reporting Items for Systematic Reviews and Meta-Analyses (PRISMA) reporting guidelines [20]. The process encompassed importing journal articles from three databases into Covidence, followed by title and abstract screening (**Figure 1**). After selecting the relevant articles, we proceeded with full-text screening and data extraction. The extracted data was then exported in the comma-separated values (CSV) file format for further analysis in Microsoft Excel, R Studio and JupyterLab.

**Figure 1.**
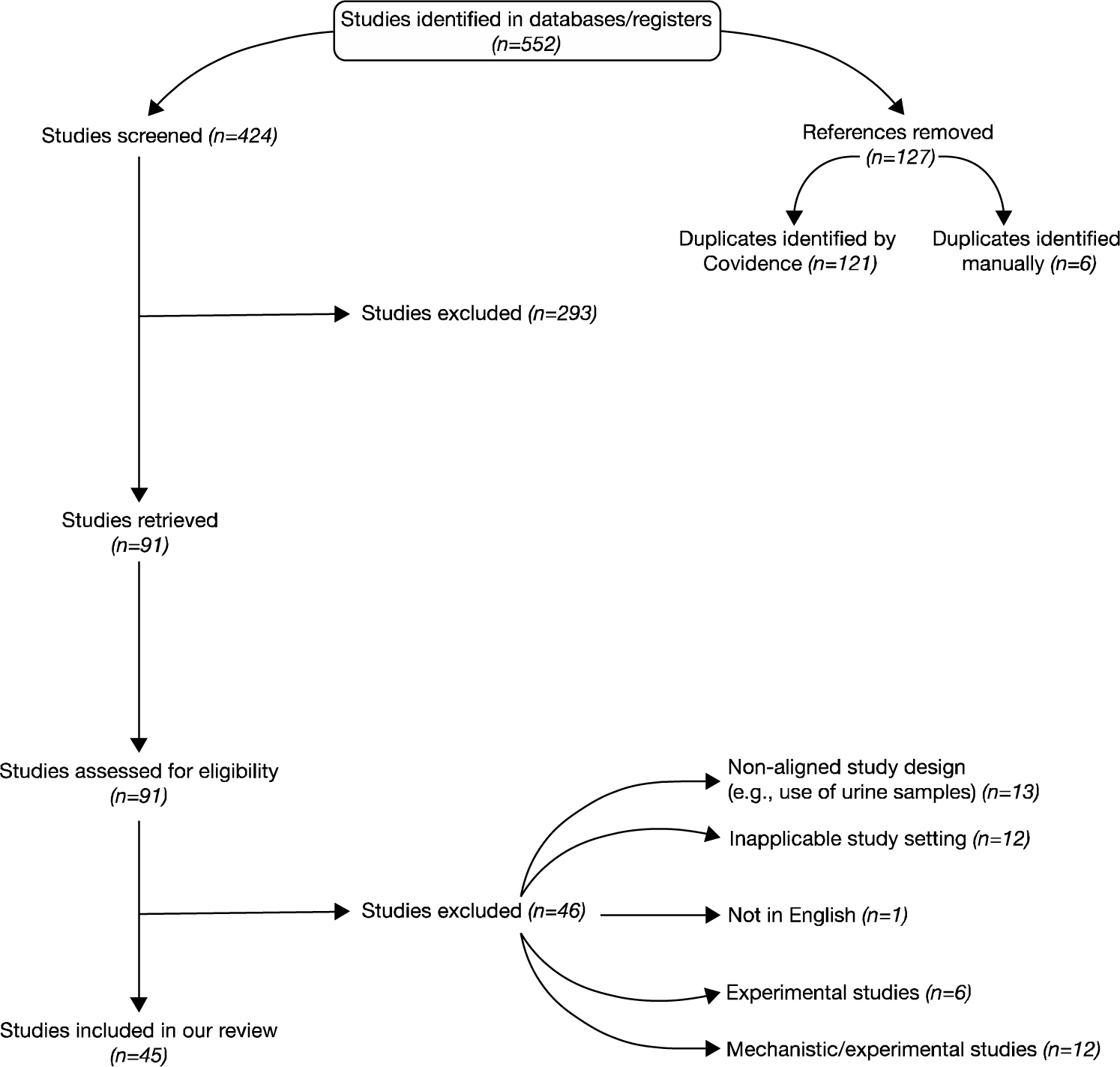
A flow diagram depicting the flow of information through the various stages of the systematic review, drawn using PRISMA.

### Generation of search terms and database selection

The search terms were derived from common terms previously associated with our topic. We reviewed the reference sections of 30 articles pertinent to the molecular epidemiology of diarrhoeagenic *E. coli*. The journals and databases in which these references were published were then noted. The selection of databases for our literature search was based on the frequency of these journals. The keywords used for the search were determined by collating those from the 30 articles mentioned above and selecting the most frequently occurring keywords related to the molecular epidemiology of diarrhoeagenic *E. coli* in Africa.

### Search strategy

Database searches were conducted using Scopus, PubMed, and EBSCOhost Research Databases. The specific strategies for each are detailed below.

#### SCOPUS

(315 results; Search date: 25^th^ April 2023)

TITLE-ABS-KEY ([“Diarrhoeagenic *Escherichia coli*”, “Verotoxigenic *Escherichia coli*”, vtec, “Verotoxigenic *E. coli*”, “Shiga Toxin-Producing *Escherichia coli*”, “Vero Cytotoxin-Producing *Escherichia coli*”, “Shiga Toxigenic *Escherichia coli*”, stec, “Shiga Toxigenic *E. coli*”, “Enteropathogenic *E. coli*”, epec, “Enteroinvasive *Escherichia coli*”, “Enteroinvasive *E. coli*”, eiec, “Diffusely Adherent *Escherichia coli*”, “Diffusely Adherent *E. coli*”, daec, “Enteroaggregative *Escherichia coli*”, eaggec, “Enteroaggregative *E. coli*”, etec, “Enterotoxigenic *E. coli*”, “Enterotoxigenic *Escherichia* coli”]) AND TITLE-ABS-KEY ([Outbreak*, Antimicrobial*, Antibiotic*, epidemic*, Pandemic*]) AND EXCLUDE (AFFILCOUNTRY, [“United States”, “United Kingdom”, “Germany”, “Canada”, “China”, “India”, “Japan”, “Brazil”, “France”, “Italy”, “Sweden”, “Spain”, “Australia”, “Iran”, “Mexico”, “Argentina”, “South Korea”, “Denmark”, “Belgium”, “Switzerland”, “Netherlands”, “Bangladesh”, “Thailand”, “Norway”, “Ireland”, “Poland”, “Indonesia”, “Finland”, “Peru”, “Turkey”, “Saudi Arabia”, “Austria”, “Pakistan”, “Chile”, “New Zealand”, “Greece”, “Hungary”, “Israel”, “Czech Republic”, “Portugal”, “Iraq”, “Viet Nam”, “Malaysia”, “Romania”, “Russian Federation”, “Taiwan”, “Nepal”, “Colombia”, “Serbia”, “Slovakia”, “Hong Kong”, “Jordan”, “Bulgaria”, “Croatia”, “Lebanon”, “Singapore”, “Uruguay”, “Bolivia”, “Slovenia”, “United Arab Emirates”, “Philippines”, “Cuba”, “Georgia”, “Laos”, “Costa Rica”, “Jamaica”, “Kuwait”, “Latvia”, “Qatar”, “Trinidad and Tobago”, “Luxembourg”, “Nicaragua”, “Venezuela”, “Cyprus”, “Ecuador”, “Estonia”, “Guatemala”, “Honduras”, “Lithuania”, “New Caledonia”, “Palestine”, “Puerto Rico”, “Yemen”, “Yugoslavia”, “Albania”, “Belarus”, “Bosnia and Herzegovina”, “Burma”, “Cambodia”, “Czechoslovakia”, “Fiji”, “Iceland”, “Kazakhstan”, “Mongolia”, “Myanmar”, “Panama”, “Paraguay”, “Saint Kitts and Nevis”, “Suriname”, “Syrian Arab Republic”]).

#### PUBMED

(225 papers; Search date, 19 April 2023) from 1977 to 2022

([“Diarrhoeagenic *Escherichia coli*[Title/Abstract]”, “VTEC[Title/Abstract]”, “Verotoxigenic *E. coli*[Title/Abstract]”, “Verotoxin-Producing *Escherichia coli*[Title/Abstract]”, “Shiga Toxin-Producing *Escherichia coli*[Title/Abstract]”, “Vero Cytotoxin-Producing *Escherichia coli*[Title/Abstract]”, “Shiga Toxigenic *Escherichia coli*[Title/Abstract]”, “STEC[Title/Abstract]”, “Shiga Toxigenic *E. coli*[Title/Abstract]”, “Enteropathogenic *E. coli*[Title/Abstract]”, “EPEC[Title/Abstract]”, “Enteroinvasive *Escherichia coli*[Title/Abstract]”, “Enteroinvasive *E. coli*[Title/Abstract]”, “EIEC[Title/Abstract]”, “Diffusely Adherent Escherichia coli[Title/Abstract]”, “Diffusely Adherent E. coli[Title/Abstract]”, “DAEC[Title/Abstract]”, “Enteroaggregative *Escherichia coli*[Title/Abstract]”, “EAggEC[Title/Abstract]”, “Enteroaggregative *E. coli*[Title/Abstract]”, “ETEC[Title/Abstract]”, “Enterotoxigenic *Escherichia coli*[Title/Abstract]”, “Enterotoxigenic *E. coli*[Title/Abstract”]) AND (Outbreak*[Title/Abstract], Antimicrobial*[Title/Abstract], Antibiotic*[Title/Abstract], Pandemic*[Title/Abstract], epidemic*[Title/Abstract]) AND Algeria[Title/Abstract], Angola[Title/Abstract], Benin[Title/Abstract], Botswana[Title/Abstract], Burkina Faso[Title/Abstract], “Burkina Faso”[Title/Abstract], Burkina Fasso[Title/Abstract], Upper Volta[Title/Abstract], “Upper Volta”[Title/Abstract], Burundi[Title/Abstract], Cameroon[Title/Abstract], Cape Verde[Title/Abstract], “Cape Verde”[Title/Abstract], Central African Republic[Title/Abstract], Chad[Title/Abstract], Comoros[Title/Abstract], “Iles Comores”[Title/Abstract], Iles Comores[Title/Abstract], Comoro Islands[Title/Abstract], “Comoro Islands”[Title/Abstract], Congo[Title/Abstract], Democratic Republic Congo[Title/Abstract], “Democratic Republic of the Congo”[Title/Abstract], Zaire[Title/Abstract], Djibouti[Title/Abstract], Egypt[Title/Abstract], Equatorial Guinea[Title/Abstract], “Equatorial Guinea”[Title/Abstract], Eritrea[Title/Abstract], Ethiopia[Title/Abstract], Gabon[Title/Abstract], Gambia[Title/Abstract], Ghana[Title/Abstract], Guinea[Title/Abstract], Guinea Bissau[Title/Abstract], “Guinea Bissau”[Title/Abstract], Ivory Coast[Title/Abstract], “Ivory Coast”[Title/Abstract], Cote d’Ivoire[Title/Abstract], “Cote d’Ivoire”[Title/Abstract], Kenya[Title/Abstract], Lesotho[Title/Abstract], Liberia[Title/Abstract], Libya[Title/Abstract], Libia[Title/Abstract], Jamahiriya[Title/Abstract], Jamahiryia[Title/Abstract], Madagascar[Title/Abstract], Malawi[Title/Abstract], Mali[Title/Abstract], Mauritania[Title/Abstract], Mauritius[Title/Abstract], Ile Maurice[Title/Abstract], “Ile Maurice”[Title/Abstract], Morocco[Title/Abstract], Mozambique[Title/Abstract], Moc_ambique[Title/Abstract], Namibia[Title/Abstract], Niger[Title/Abstract], Nigeria[Title/Abstract], Rwanda[Title/Abstract], Sao Tome[Title/Abstract], “Sao Tome”[Title/Abstract], Senegal[Title/Abstract], Seychelles[Title/Abstract], Sierra Leone[Title/Abstract], “Sierra Leone”[Title/Abstract], Somalia[Title/Abstract], South Africa[Title/Abstract], “South Africa”[Title/Abstract], Sudan[Title/Abstract], South Sudan[Title/Abstract], “South Sudan”[Title/Abstract], Swaziland[Title/Abstract], Tanzania[Title/Abstract], Tanganyika[Title/Abstract], Zanzibar[Title/Abstract], Togo[Title/Abstract], Tunisia[Title/Abstract], Uganda[Title/Abstract], Western Sahara[Title/Abstract], “Western Sahara”[Title/Abstract], Zambia[Title/Abstract], Zimbabwe[Title/Abstract], Africa[Title/Abstract], Africa*[Title/Abstract], Southern Africa[Title/Abstract], West Africa[Title/Abstract], Western Africa[Title/Abstract], Eastern Africa[Title/Abstract], East Africa[Title/Abstract], North Africa[Title/Abstract], Northern Africa[Title/Abstract], Central Africa[Title/Abstract], Sub Saharan Africa[Title/Abstract], Subsaharan Africa[Title/Abstract], Sub-Saharan Africa[Title/Abstract]).

#### EBSCO HOST

via EBSCOhost Research Databases (176 papers; Search date, 24 April 2023): Using Africa-Wide Information and CINAHL

AB ([“Diarrhoeagenic *Escherichia coli*”, “VTEC”, “Verotoxigenic *E. coli*”, “Verotoxin-Producing *Escherichia coli*”, “Shiga Toxin-Producing *Escherichia coli*”, “Vero Cytotoxin-Producing *Escherichia coli*”, “Shiga Toxigenic *Escherichia coli*”, “STEC”, “Shiga Toxigenic *E. coli*”, “Enteropathogenic *E. coli*”, “EPEC”, “Enteroinvasive *Escherichia coli*”, “Enteroinvasive *E. coli*”, “EIEC”, “Diffusely Adherent *Escherichia coli*”, “Diffusely Adherent *E. coli*”, “DAEC”, “ Enteroaggregative *Escherichia coli*”, “EAggEC”, “Enteroaggregative *E. coli*”, “ETEC”, “Enterotoxigenic *E. coli*”, “Enterotoxigenic *Escherichia coli*”]) AND AB([Outbreak*, Antimicrobial*, Antibiotic*, Pandemic*, Epidemic*]) AND AB ([“Algeria”, “Angola”, “Benin”, “Botswana”, “Burkina Faso”, “Burkina Faso”, “Burkina Fasso”, “Upper Volta”, “Upper Volta”, “Burundi”, “Cameroon”, “Cape Verde”, “Cape Verde”, “Central African Republic”, “Chad”, “Comoros”, “Iles Comores”, “Comoro Islands”, “Congo”, “Democratic Republic Congo”, “Democratic Republic of the Congo”, “Zaire”, “Djibouti”, “Egypt”, “Equatorial Guinea”, “Eritrea”, “Ethiopia”, “Gabon”, “Gambia”, “Ghana”, “Guinea”, “Guinea Bissau”, “Guinea Bissau”, “Ivory Coast”, “Cote d’Ivoire”, “Kenya”, “Lesotho”, “Liberia”, “Libya”, “Libia”, “Jamahiriya”, “Jamahiryia”, “Madagascar”, “Malawi”, “Mali”, “Mauritania”, “Mauritius”, “Ile Maurice”, “Ile Maurice”, “Morocco”, “Mozambique”, “Moçambique”, “Namibia”, “Niger”, “Nigeria”, “Rwanda”, “Sao Tome”, “Sao Tome”, “Senegal”, “Seychelles”, “Sierra Leone”, “Sierra Leone”, “Somalia”, “South Africa”, “South Africa”, “Sudan”, “South Sudan”, “South Sudan”, “Swaziland”, “Tanzania”, “Tanganyika”, “Zanzibar”, “Togo”, “Tunisia”, “Uganda”, “Western Sahara”, “Western Sahara”, “Zambia”, “Zimbabwe”, “Africa”, “Africa*”, “Southern Africa”, “West Africa”, “Western Africa”, “Eastern Africa”, “East Africa”, “North Africa”, “Northern Africa”, “Central Africa”, “Sub Saharan Africa”, “Subsaharan Africa”, “Sub-Saharan Africa”]).

### Study Eligibility Criteria

We incorporated studies that specifically reported on the prevalence and or antimicrobial resistance patterns of diarrhoeagenic *E. coli* pathotypes derived from human sources but excluded samples from extra-intestinal sources and those that did not use molecular methods to confirm the pathotype.

Studies were excluded at the screening and full-text review stages if they were systematic or literature reviews, they exhibited an unclear study design, the articles were not written in English, or they sourced *E. coli* from non-human origins such as water, animals, soil, or food. Studies based on regions outside Africa (e.g., Europe, Asia, Americas, Australasia) were similarly excluded, along with papers reporting drug or mechanistic trials.

### Study Quality Assessment

Each study’s quality was scrutinised by two independent reviewers using a designated quality assessment protocol. The laboratory methodologies implemented needed to align with global recommended standards. These methods should have been confirmatory rather than mere screening tools.

### Title and Abstract Screening

Preliminary screening of the gathered studies was done based on their titles and abstracts. Two reviewers determined the eligibility of each study for inclusion. In cases where the reviewers’ decisions clashed, a consensus was reached through discussion.

### Full-Text Screening

At this juncture, the complete text of each article was meticulously perused by two reviewers to gauge its relevance. Any disagreement between the reviewers was settled through a mutual discussion to reach a final decision.

### Data Extraction Strategy

We employed the Covidence software to devise a data extraction protocol tailored to accrue pertinent data about the antimicrobial resistance and prevalence of diarrhoeagenic *E. coli* pathotypes across Africa. This protocol was formulated and refined with the insights of four reviewers until a unanimous consensus was reached. During the extraction phase, each study was critically examined by two reviewers. Discrepancies in data extracted by the reviewers were addressed and resolved by a third reviewer’s intervention.

### Data Analysis

Upon the completion of data extraction, the results were transitioned into a comma-separated value (CSV) file format and integrated into Microsoft Excel for subsequent analysis. Statistical computations were predominantly executed using Python v3.10.9 via the JupyterLab interactive development environment v3.5.3. Data on pathotype prevalence was extracted and processed using Python’s pandas library v2.0.3. The processed data, detailing the number of cases and the sample size for each study, was then passed to the R statistical environment for further analysis.

Using R’s metafor package v4.2-0, a DerSimonian and Laird random-effects meta-analysis was performed. The metafor package computes effect sizes and associated variances, facilitating meta-analytic pooling of prevalence rates across studies. For each study, the point estimate (proportion) of prevalence and its variance were computed using the escalc function, which utilises the proportion of cases (xi) over the sample size (ni) with the “PFT” measure.

Subsequently, the rma function from the metafor package was employed to compute the pooled random-effects estimate while considering between-study heterogeneity. This meta-analysis yielded effect sizes (or meta-estimates) for each study and an overall pooled effect size representing the cumulative prevalence estimate.

Pairwise comparisons were conducted to compare the prevalence estimates of various diarrhoeagenic *E. coli* pathotypes. The absolute differences in prevalence estimates between each pair of pathotypes were computed. To ascertain the significance of these differences, p-values were derived by comparing these differences to a normal distribution. Given the multiple comparisons, the Bonferroni correction was applied to adjust the significance level, ensuring the control of the family-wise error rate. A difference was deemed statistically significant if its associated p-value was lower than the Bonferroni-adjusted significance threshold.

Python library matplotlib v3.7.2 was used to generate a forest plot, displaying the prevalence rate for each study, along with the 95% confidence intervals. The overall pooled prevalence rate was distinctly highlighted to emphasise the aggregate findings of the analysis.

For data on antimicrobial resistance, the frequency and percentage of non-susceptible isolates for each antibiotic class were documented for studies where antibiograms were reported. For a selection of antibiotics, we used Stata v17 statistical (StataCorp) software to carry out a meta-analysis to determine the pooled resistance at the pathotype level.

We employed the Chi-Square and Fisher’s Exact tests to investigate the statistical significance of observed antibiotic nonsusceptibility across different antibiotic classes. Each antibiotic class’ observed frequencies were compared to expected frequencies based on the assumption of even distribution within that class. Specifically, when any expected frequency count in a class was less than or equal to five, the Fisher’s Exact test was employed; this was especially pertinent for 2×2 tables but was extended here through a series of 2×2 tables, with the smallest p-value taken as representative. The Chi-Square test was employed in cases where all expected frequencies were above five. A p-value less than 0.05 was considered indicative of a significant departure from the expected distribution, thus suggesting that the observed frequencies were unlikely due to random variation alone.

### Data Visualisation

All visualisations were created using Python’s matplotlib library v3.7.2. The Set3 palette from seaborn library v0.12.2 was employed to ensure that distinct categories (like pathotypes in our study) were easily distinguishable. For the distribution of pathotype-specific studies by country, a heatmap was used, where each cell in the heatmap displays the number of studies, with the colour intensity indicative of the quantity (the darker the shade, the higher the number). The methods used in the reviewed studies were visualised using a stacked bar chart, with the colour of each bar segment signifying a distinct method used in the studies in percentage terms. For the meta-analysis results, a forest plot-like visualisation was employed, where the estimated prevalence from different studies, along with their 95% confidence intervals, were presented using error bars. This format allowed for a clear comparison of prevalence estimates across studies and pathotypes.

## Results

### Geographical distribution

Forty-five publications were reviewed for data extraction, spanning 18 African countries (**Supplementary Table 1**). Most of the studies emerged from Kenya (24%) and South Africa (18%). Of the 45 articles, 76% (34/45) reported on EPEC, 69% (31/45) reported on EAEC and ETEC, 44% (20/45) reported on EIEC, 36% (16/45) reported on STEC/VTEC, 11% (5/45) reported on DAEC, and only 6% (3/45) reported hybrid strains.

An interesting observation from the geographical data was the pronounced concentration of reports of specific pathotypes in certain regions. Kenya, South Africa, and Nigeria emerged as areas where EPEC, EAEC, and ETEC studies commonly emanated — a pattern clearly illustrated in **Figure 2A**.

**Figure 2.**
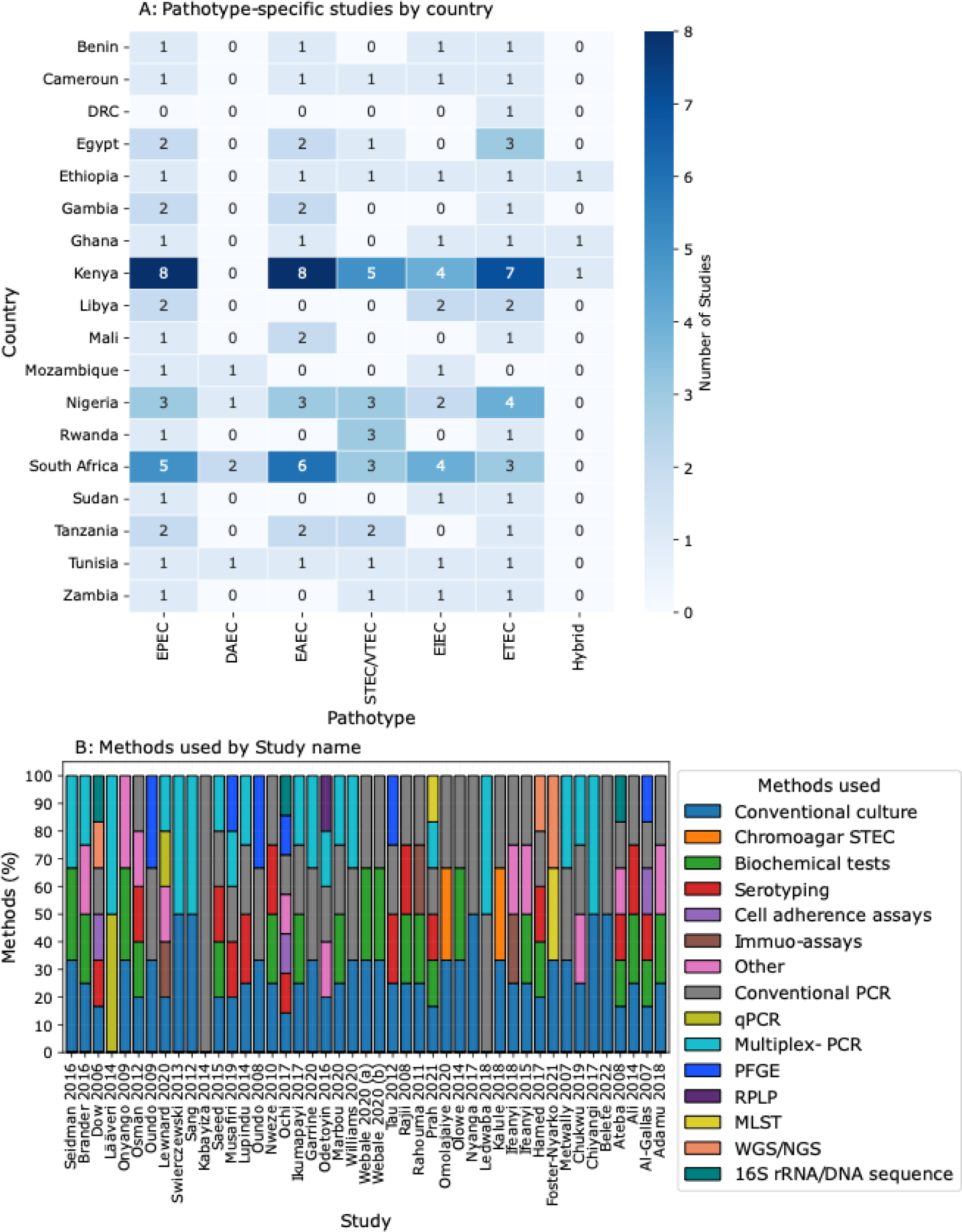
Overview of pathotype-specific studies by country and methodologies employed. **Panel A** (Pathotype-specific studies by country) illustrates the distribution of pathotype-specific studies across various countries. The countries are displayed on the y-axis, while distinct pathotypes are on the x-axis. The colour intensity, progressing from light to dark, represents an increasing number of studies, with the precise number annotated within each cell. **Panel B** (Methods used) details the techniques used across the studies reviewed. The methods are enumerated along the x-axis; each colour in the bars corresponds to a distinct method utilised in the research, as indicated by the colour-coded legend. The percentage (on the y-axis) denotes the prevalence of each method in each study. PFGE, Pulsed Field Gel Electrophoresis; MLST, Multilocus Sequencing Typing; WGS, Whole Genomics Sequencing; NGS, Next generation sequencing; PCR, Polymerase Chain Reaction; qPCR, quantitative Polymerase Chain Reaction; RFLP, Restriction Fragment Length Polymorphism; Other, includes techniques like haemolytic activities, verotoxicity tests, ELISA, transconjugation assays and the Colilert test.

### Study types

Most studies (43/45, 96%) comprised reports from non-outbreak settings, while only 4% (2/45) represented the retrospective use of outbreak samples. At the same time, 69% (31/45) were cross-sectional and 27% (12/45) were classified as case-control studies.

### Population characteristics in the reviewed studies

The human population samples covered a broad age spectrum. Still, the focus was predominantly on younger children despite many publications addressing more than one age group. One of the publications reported on neonates under 28 days, 31% (14/45) on infants under five years and 58% (26/45) on children under 18. Fewer reports (7/45, 16%) focused on adults over 18 years, including one focusing on older people over 65 years, and 27% (12/45) did not specify the age groups.

In terms of specific population categories, 4% (2 out of 45) were in rural settings, 7% (3 out of 45) involved food handlers, and 2% (1 out of 45) each came from urban and peri-urban areas, with or without livestock. The rest were from unique reports, such as travellers’ diarrhoea, low-income populations, a wedding party, and those suffering from underlying diseases.

### Study sites

All the examined publications reported on stool samples primarily collected in hospital settings at 67% (30 out of 45). In contrast, 13% (6 out of 45) of the studies collected samples from both hospital and community settings, and 16% (7 out of 45) were from community settings alone.

Of the 45 studies, all (100%) reported patients presenting with diarrhoea. Of these, 40% (18 out of 45) reported patients displaying severe signs of diarrhoea, including bloody diarrhoea (10%), vomiting (14%), and fever (14%).

### Diagnostic Laboratory Techniques Utilised from the African sourced publications

We explored the methodologies employed to detect diarrhoeagenic *E. coli*. From the 45 publications scrutinised, more than 15 analytic tools were identified (**Figure 2B**). Most researchers (42/45; 93%) began their investigations with conventional culture techniques to isolate pathogens from clinical specimens. Subsequent screening and verification utilised a variety of approaches, including biochemical identification methods, conventional or multiplex PCR, and serotyping. Notably, only a few studies (3/45, 7%) employed sequencing tools, underscoring the limited capacity for advanced sequencing tools in foodborne research on the continent. Researchers often sought collaboration with national or international laboratories when specific tools were unavailable.

### Prevalence of Diarrhoeagenic *E. coli* Pathotypes

Given that the reviewed publications did not consistently encompass all six main diarrhoeagenic pathotypes, our calculations for individual pathotypes included only those explicitly reported. Consequently, studies that investigated specific pathotypes did not contribute prevalence data for other types that were not within their research scope. Our analysis highlighted EAEC as the most prevalent pathotype (43%; 95% CI, 30% – 55%) (**Figure 3A**), while STEC (**Figure 3E**) and EIEC (**Figure 3F**) presented the lowest prevalence at 28% (95%CI, 14% – 42%) and 24% (95%CI, 17% – 32%), respectively. Furthermore, ETEC, DAEC, and EPEC—inclusive of atypical EPEC— also emerged as notably prevalent pathotypes with prevalence rates of 36% (95% CI, 27% – 45%), 36% (95% CI, 16% – 57%), and 31% (95% CI, 21% – 40%), respectively (**Figures 3B – 3D**). Notably, there were only three reports of hybrid strains throughout the studies under review.

**Figure 3.**
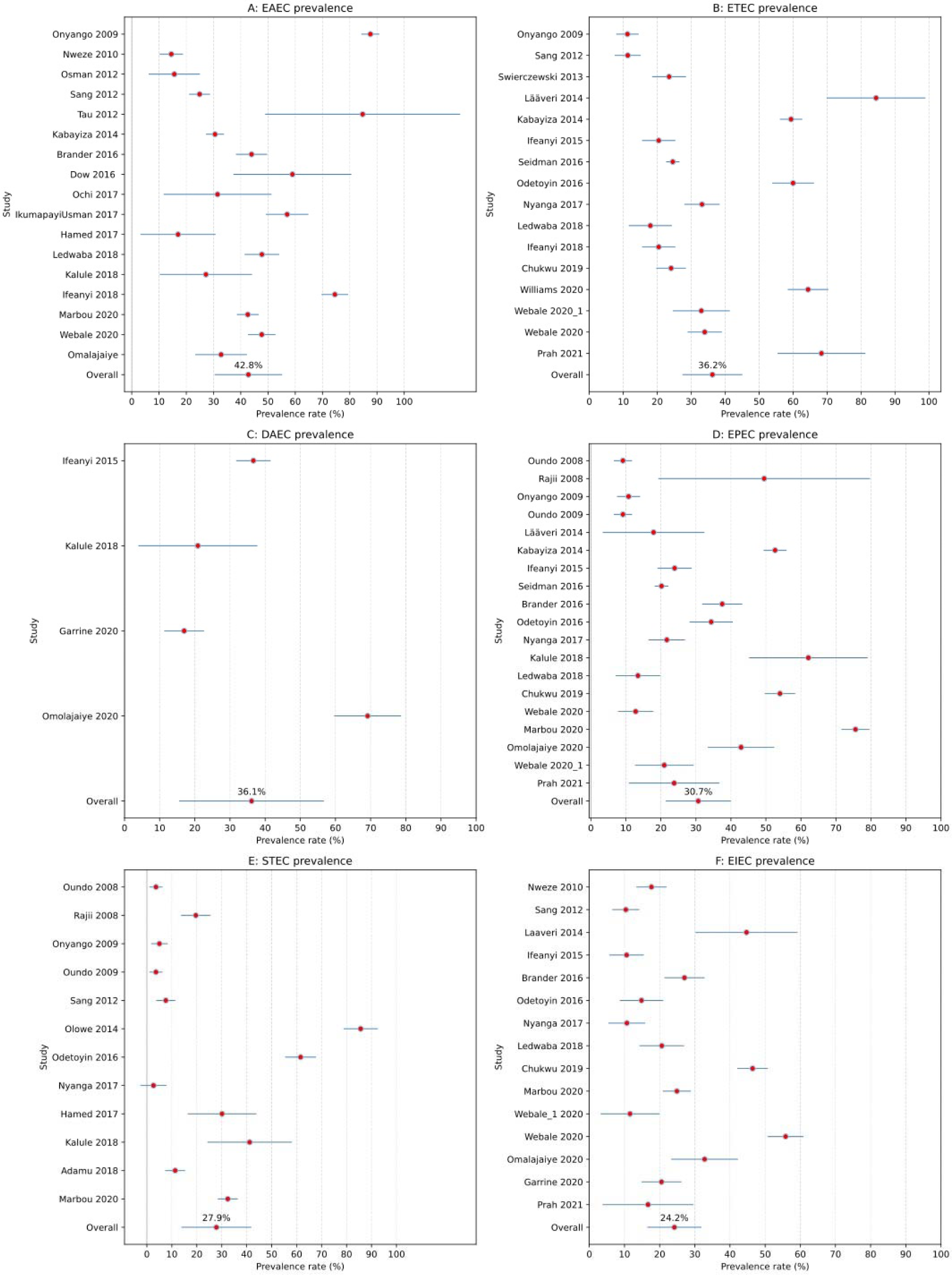
Prevalence of enteropathogenic bacterial pathotypes. The figure displays six distinct forest plots, each highlighting the prevalence of a specific enteropathogenic bacterial pathotype. Red circles represent the estimated prevalence rates from individual studies. Accompanying these markers, horizontal blue lines illustrate the 95% confidence interval (CI) for each respective rate. Each panel is dedicated to a different bacterial pathotype. Panel A elucidates the prevalence of Enteroaggregative *E. coli* (EAEC), while Panel B depicts Enterotoxigenic *E. coli* (ETEC). Similarly, Panels C through F focus on Diffusely adherent *E. coli* (DAEC), Enteropathogenic *E. coli* (EPEC), Shiga toxin-producing *E. coli* (STEC), and Enteroinvasive *E. coli* (EIEC), respectively. The percentage value annotated above the ’Overall’ marker indicates the cumulative meta-analysed prevalence rate associated with each pathotype. As a point of reference, a vertical grey line is drawn at the 0% prevalence rate mark, and dashed gridlines are included at regular intervals to facilitate a clearer understanding of the prevalence percentages.

Comparative analyses of the prevalence of different pathotypes highlighted significant disparities among them (**Supplementary Table 2**). EAEC exhibited the highest prevalence and was significantly more prevalent than ETEC, DAEC, EPEC, STEC, and EIEC, with differences in prevalence estimates ranging from approximately 6.56% to 18.61%. ETEC’s prevalence was notably different from that of EPEC, STEC, and EIEC, though not significantly different from DAEC. Furthermore, DAEC showed a significantly distinct prevalence from EPEC, STEC, and EIEC. EPEC and STEC differed insignificantly in prevalence. In contrast, there were noticeable differences between the prevalence of EPEC and EIEC and between STEC and EIEC.

### Commonly Used Susceptibility Testing Techniques and Interpretive Standards

In our analysis of methodologies employed for assessing and interpreting antibiotic resistance across the selected studies, the Clinical and Laboratory Standards Institute (CLSI) guidelines emerged as the preferred framework. A substantial 90% (27/30) of the studies that detailed antibiotic resistance determinations opted for the CLSI guidelines. On the other hand, the EUCAST guidelines found favour in only 10% (3/30) of the publications, with a number inclusive of reports following the directives of the Antibiogram Committee of the French Microbiological Society.

Regarding the antimicrobial susceptibility testing approach, the Kirby-Bauer disc diffusion method was the dominant technique, which was utilised by 73% (22/30) of the publications that reported non-susceptibility to an antibiotic. Another 23% (7/30) integrated both the disc diffusion and micro-broth dilution methods. A minority, 10% (3/30), relied solely on the micro-broth dilution method. There was a solitary report of the VITEK system in use, with results closely mirroring those derived via the micro-broth dilution method.

Notably, only seven studies explored antibiotic-resistant genes, either as an exclusive method or in conjunction with the susceptibility assessment techniques mentioned earlier.

### Antibiotic Resistance Among Diarrhoeagenic *E. coli*

Of 30 studies presenting antimicrobial resistance outcomes, a cumulative 602 antimicrobial resistance testing outcomes could be classified as antibiotic “non-susceptible”, comprising 87% (n=521) resistant and 14% (n=81) intermediate resistant isolates (**Table 1**).

**Table 1:** Prevalent rates (counts) of non-susceptibility (intermediate and resistant phenotypes) to commonly used antibiotics.

| Antibiotic class | Count of non-susceptibility by antibiotic | Frequency (by class) | Percent | Cumulative | Test used | p-value |
| --- | --- | --- | --- | --- | --- | --- |
| Quinolones | Nalidixic acid, 32 (30.48%); Ciprofloxacin, 52 (49.52%); Norfloxacin, 20 (19.05%); Enrofloxacin, 1 (0.95%) | 105 | 17.44 | 74.92 | Chi-Square | 2.5911E-11 |
| Cephems | Ceftriaxone, 26 (26.53%); Cefotaxime, 20 (20.41%); Ceftazidime, 20 (20.41%); Cefpodoxime, 5 (5.10%); Cephalexin, 1 (1.02%); Cefuroxime, 14 (14.29%); Cephazolin, 5 (5.10%); Cefoxitin, 3 (3.06%); Cefepime, 2 (2.04%) | 98 | 16.28 | 38.7 | Chi-Square | 1.9844E-13 |
| Aminoglycosides | Gentamycin, 39 (49.37%); Streptomycin, 22 (27.85%); Kanamycin, 8 (10.13%); Neomycin, 1 (1.27%); Amikacin, 8 (10.13); Tobramycin, 1 (1.27%) | 79 | 13.12 | 51.83 | Chi-Square | 1.8365E-16 |
| Penicillins | Ampicillin, 61 (80.26%); Ticarcillin, 1 (1.32%); Ofloxacin, 12 (15.79); Amoxicillin, 2 (2.63%) | 76 | 12.62 | 12.62 | Chi-Square | 1.7065E-27 |
| Folate pathway inhibitors | Trimethoprim, 14 (23.73%); Trimethoprim + sulfamethoxazole, 35 (59.32%); Sulphamethoxazole, 10 (16.95%) | 59 | 9.8 | 22.43 | Chi-Square | 0.00010417 |
| Phenicol | Chloramphenicol, 46 (100%) | 46 | 7.64 | 96.67 | Chi-Square |  |
| Tetracyclines | Tetracycline, 38 (95%); Oxycycline, 1 (2.50%); Doxycycline, 1 (2.50%) | 40 | 6.64 | 89.03 | Chi-Square | 1.3686E-15 |
| Macrolides | Erythromycin, 19 (55.88%); Clarithromycin, 5 (14.71%); Azithromycin, 10 (29.41%) | 34 | 5.65 | 57.48 | Chi-Square | 0.01178207 |
| β-lactam-inhibitor combinations | Amoxicillin-clavulanic acid, 23 (92.0%); Piperacillin-tazobactam, 2 (8.00%) | 25 | 4.15 | 79.07 | Chi-Square | 2.6691E-05 |
| Carbapenem | Ertapenem, 1 (5%); Meropenem, 11 (55%); Imipenem, 8 (40%) | 20 | 3.32 | 82.39 | Chi-Square | 0.0192547 |
| Lipopeptides | Colistin sulfate, 7 (100%) | 7 | 1.16 | 98.33 | Chi-Square |  |
| Glycylcycline | Tigecycline, 6 (100%) | 6 | 1 | 99.34 | Chi-Square |  |
| ESBL | ESBL, 4 (100%) | 4 | 0.66 | 100 | Fisher's Test | 1 |
| Nitrofurans | Nitrofurantoin, 3 (100.00%) | 3 | 0.5 | 97.17 | Fisher's Test | 1 |

Among these, Quinolones surfaced as the most frequently encountered resistant class (p-value, 2.59E-11), with a frequency of 105. Following closely were the Cephems, registering a frequency of 98 (p-value, 1.98E-13), then Aminoglycosides and Penicillins, with frequencies of 79 and 76, respectively. Folate pathway inhibitors followed with a frequency of 59 (p-value, 0.0001).

Phenicols and Tetracyclines came next, with frequencies of 46 and 40, respectively. Non-susceptibility to Macrolides, ß-lactam-inhibitor combinations, and Carbapenem classes were also observed. Notably, only four instances of ESBL phenotypes were observed (**Table 1**).

Our analysis revealed variable resistance patterns among the different *E. coli* pathotypes for the studies that reported antimicrobial susceptibility (AST) data (**Supplementary Figures 1 – 5**). Despite a prevalence of 36% for DAEC, none of the studies that reported on this pathotype, namely Garrine 2020, Omolajaiye 2020, Kalule 2018, and Ifeanyi 2015, provided data regarding antimicrobial susceptibility testing (ASTs). Hence, no available data exists on the prevalence of resistance among DAEC isolates. By contrast, STEC, ETEC, and EPEC isolates exhibited strikingly high resistance to ampicillin (the most frequently reported antibiotic among the reviewed studies) with prevalence rates and 95% confidence intervals (CIs) of 72% (13% – 100%), 73% (58% – 89%), and 72% (46% – 98%), respectively (**Supplementary Figures 1 – 3**), albeit with considerably wide confidence intervals, hinting at the variability within the data and the small number of studies reporting AST data. On the other hand, pooled estimates for ampicillin resistance among EAEC and EIEC were 43.3% (95% CI: 40% – 47%) and 43% (95% CI: 36% – 51%), respectively. When assessing antimicrobial susceptibility across all diarrhoeagenic *E. coli* without distinguishing between individual pathotypes, the pooled resistance was as follows: 73% (95% CI: 64% – 83%) for ampicillin, 32% (95% CI: 19% – 46%) for gentamicin, 22% (95% CI: 12% – 33%) for nalidixic acid, 14% (95% CI: 8% – 20%) for ciprofloxacin, and 14% (95% CI: 3% – 25%) for ceftriaxone (**Figure 4**).

**Figure 4:**
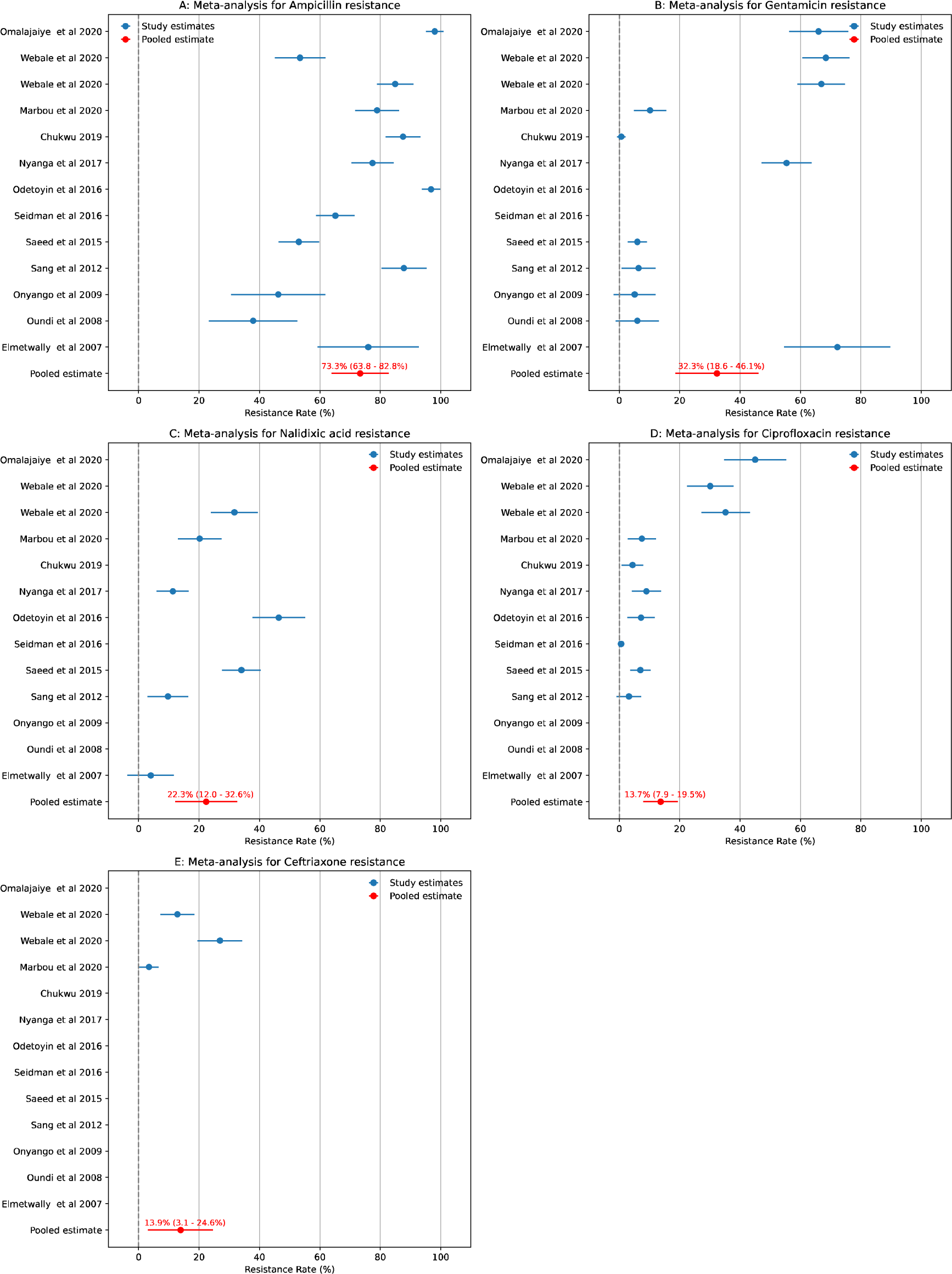
Meta-analysis of antibiotic resistance prevalence in diarrhoeagenic *E. coli* across studies. This figure showcases a series of subplots, each dedicated to representing the prevalence of resistance to a particular antibiotic among all diarrheagenic *Escherichia coli* (DEC) strains. Each blue dot pertains to a specific study and displays the proportion of DEC samples in that study that exhibited resistance to the antibiotic under consideration. The horizontal position of each dot indicates the resistance percentage. The vertical position denotes the study from which the data originates. The error bars, extending from each dot, represent the 95% confidence interval (CI) of the resistance percentage for that study. The red dot in the first position of each subplot represents the pooled estimate of the resistance rate from a random effects model. The horizontal lines connected to the red dot indicate the 95% CIs of the pooled estimate. Beside each pooled estimate dot is a label specifying the exact resistance percentage and the corresponding 95% CIs.

## Discussion

This study reviewed the prevalence and antimicrobial resistance patterns of diarrhoeagenic *E. coli* pathotypes in Africa. A significant proportion of reported studies emanated from Kenya, South Africa, and Nigeria. However, these numbers may not accurately reflect the actual disease prevalence across the continent. The disparity in the number of diarrhoeagenic *E. coli* studies underscores the varying diagnostic and surveillance practices across African nations. This can be attributed to the more developed healthcare infrastructures in Kenya, South Africa, and Nigeria [21], emphasising the need for enhanced surveillance and pathotype-specific interventions across Africa.

Our review showed that EAEC and ETEC were the most common diarrhoeagenic *E. coli* pathotypes among the extant literature, with EIEC being the least prevalent. This finding aligns with the conclusions of the Global Enteric Multicenter Study (GEMS), where ETEC, among others, was a significant contributor to moderate-to-severe diarrhoea in young children. Earlier studies have highlighted EAEC and ETEC as substantial contributors to childhood diarrhoea [1]. However, our review discerned a higher prevalence of ETEC, EAEC, and DAEC. In line with earlier research, we identified an underreporting of EHEC, likely due to its overshadowing by other more readily detectable pathogens that cause dysentery [22].

Interestingly, only two studies pinpointed outbreaks triggered by a specific diarrhoeagenic *E. coli* pathotype, with one being attributed to a novel heat-stable enterotoxin-producing strain of Enteroaggregative *E. coli* [23]. The evident lack of real-time surveillance for foodborne pathogens in many regions likely obscures the detection and actual frequency of outbreaks. Moreover, most studies in our review opted for culture-based diagnostic methods despite the evolution and optimisation of more sensitive genomic epidemiology techniques that could be adapted to low-resource settings [14, 15, 24, 25].

A notable observation from our study is the marked scarcity of research concerning hybrid strains. Hybrid strains often manifest with more severe clinical outcomes than their non-hybrid counterparts, as emphasised by Santos et al. [26]. One plausible explanation for the limited detection of these strains could be the lack of genomic capacity in the region to identify specific genotypic markers indicative of hybridity. This is further corroborated by the limited number of studies incorporating genomics into their *E. coli* pathotype surveillance within the reviewed literature. Consequently, there’s a compelling argument for the broader integration of genomics in Africa’s diagnostic landscape. Doing so will not only enhance the identification of such hybrid strains but also augment the continent’s capacity to anticipate and mitigate potential outbreaks.

Moreover, our analysis unveiled a trend of heightened resistance to pivotal antibiotics, notably Quinolones and Cephalosporins. Due to the fragmented nature of the reports, we could not determine absolute prevalence rates of resistance for diarrhoeagenic *E. coli.* While our results provide important insights, it’s crucial to note that this data only presents a snapshot of the situation, as they are based on a limited number of studies. We need dedicated and meticulously designed epidemiological studies to grasp the true prevalence of resistance. A comprehensive approach, grounded in surveillance and well-structured research, is paramount to understanding and combating the rising antimicrobial resistance tide. However, our findings warrant continuously enhanced efforts to prevent the increase and emergence of antimicrobial resistance on the continent [27].

In a silver lining, a marked low resistance was observed to extended-spectrum beta-lactamases (ESBLs) and third-generation cephalosporins. While this can be construed as a positive indication that the continent might be relatively shielded from the burgeoning global ESBL challenge, caution is still warranted. The low prevalence of ESBLs could reflect the limited number of studies that actively sought out ESBL determinants or harnessed genomics to characterise resistance against this antibiotic class. Given the potential clinical implications, it becomes imperative for Africa to sustain vigilant monitoring in this domain, ensuring that the continent remains a step ahead in the battle against antimicrobial resistance.

Of the four studies that documented ESBL production among diarrhoeagenic *E. coli,* one reported on the environmental correlates of antimicrobial resistance and noted that children whose caregivers used a shared pit latrine or who openly defecated were more likely to carry multidrug-resistant bacteria than those with flush or unshared toilets [36]. This underscores the need for broader, community-based research on foodborne pathogens. Unfortunately, our review noted that most studies reported on children less than 18 years of age during a health crisis and that more than two-thirds of the samples studied were reported from hospital sites. However, the actual burden of the disease within a community is better represented by community samples.

The interplay between antibiotic resistance and the virulence of pathogenic bacteria, including *E. coli,* has garnered significant global concern [1, 28, 29]. While some previous studies have suggested reduced virulence among multidrug resistant *E. coli* isolates relative to sensitive strains [30], others have emphasised that the acquisition of antimicrobial resistance does not necessarily compromise microbial fitness [31].

Consistent with this notion, epidemiological data indicate that antibiotic resistance and virulence factor carriage are linked in *E. coli* populations in some community settings [32]. A related study showed that the expression of virulence factors led to the formation of an antibiotic-tolerant subpopulation [33] and that antibiotic treatment indeed may select for virulence [34]. In addition to drug resistance, treatment failure on the use of antibiotics in a clinical setting could be due to tolerance and or persistence to antibiotics [35]. Importantly, community-based surveillance studies are pivotal, as evidenced by findings linking sanitation practices with antibiotic resistance patterns [36]. However, our review discerned a focus on younger populations and hospital-based studies, underscoring the need for broader, community-based research.

Accurate pathotype identification remains a challenge due to the complexities associated with conventional laboratory techniques. The predominant reliance on the disk diffusion method over minimum inhibition concentration (MIC) for antimicrobial testing introduces further complexity due to varying sensitivity levels. Notwithstanding these challenges, a noticeable trend toward employing more sensitive diagnostic methodologies has emerged, suggesting an optimistic trajectory for future African studies.

On this note, the recent endeavours by Africa CDC, particularly through the foodborne disease focus group, underscore the continent’s readiness to embrace advanced surveillance platforms for tracking foodborne disease outbreaks. Leveraging high-resolution techniques incorporating genomics, such as whole-genome sequencing (WGS), will not only elevate the precision of outbreak detection but also revolutionise our understanding of disease spread and antimicrobial resistance patterns. This approach, if widely adopted, positions Africa at the forefront of combating foodborne pathogens and ensuring the health and safety of its populace.

In our systematic review of published data on diarrhoeagenic *E. coli* from the African continent’s public health sector, we encountered significant challenges in data collation. The heterogeneity in study designs and methodologies resulted in fragmented outputs. Notably, a limited number of studies reported AST data, and where they occurred, antimicrobial resistance profile determinations utilised a diverse array of antibiotics. Furthermore, many studies reported concentrations not aligned with the CLSI or EUCAST breakpoint guidelines. This disparity underscores the pressing need for standardised testing, reporting, and interpretive guidelines tailored to Africa’s unique demographics, geographies, and economic scales. Such standardisation would ensure reproducibility and establish a robust platform for historic surveillance, enabling timely assessment of risks to the healthcare sector across the continent.

### Limitations

This review was meticulously designed with comprehensive search criteria to encompass a broad spectrum of studies focusing on *E. coli* pathotypes. Nevertheless, given the vast expanse of scientific literature on the topic, there remains a possibility that some pertinent studies might have been inadvertently overlooked.

Furthermore, significant heterogeneity was observed among the studies reviewed, stemming from differences in study design, population demographics, geographical location, and methods of pathotype identification. Such heterogeneity can invariably influence the overall prevalence estimates. While rigorous meta-analytic techniques were employed to mitigate this variation, it’s crucial to acknowledge that the reported rates may not capture the complete picture. They might, in fact, reflect under-reporting due to a variety of reasons, including limited diagnostic capacities or logistical constraints in specific settings.

### Conclusions

Our comprehensive review of published data on diarrhoeagenic *E. coli* from the African continent underscores the significant heterogeneity in study designs, methodologies, population characteristics, and sample collection sites. Kenya, South Africa, and Nigeria emerge as hotspots for research into particular pathotypes, with a noticeable focus on EPEC, EAEC, and ETEC. The propensity for hospital-based sample collection is evident, with a notable divergence in methodologies employed for both detection and antibiotic resistance assessments.

EAEC’s prevalence as the dominant pathotype, juxtaposed with the striking low reports of hybrid strains, underlines the need for targeted surveillance and management strategies. The alarmingly high resistance rates to commonly used antibiotics, including emerging resistance to crucial drugs such as ciprofloxacin, underscore the imminent threat of antibiotic resistance in the region. This calls for urgent action in the form of robust antibiotic stewardship programs, harmonised surveillance efforts, and educational campaigns aimed at healthcare providers and the public.

Our findings elucidate the dominance of the CLSI guidelines in antibiotic resistance determination, indicating a potential avenue for standardisation and consolidation of antimicrobial resistance reporting. The prominence of the Kirby-Bauer disc diffusion method highlights the method’s accessibility and utility in the region.

In light of these insights, there is a pronounced need for the African continent to foster standardised testing, reporting, and interpretative guidelines tailored to its unique demographic and geographic contexts. This will be instrumental in optimising diagnostics, treatment protocols, and mitigation strategies against the looming threat of antimicrobial-resistant diarrhoeagenic *E. coli* and other pathogens in the region.

## Data Availability

All data produced in the present work are contained in the manuscript

## Acknowledgements

The systematic review, writing workshops and in-person meetings for the FBD expert group members were funded by the Bill & Melinda Gates Foundation grant INV-033857. The authors wish to express their profound gratitude to the Africa PGI of Africa CDC for their unwavering support throughout this systematic review. Additionally, we extend our heartfelt thanks to Professor Kathryn E. Holt for dedicating her valuable time and providing insightful feedback on the draft manuscript.

## Supplementary figures

**Supplementary Figure 1:**
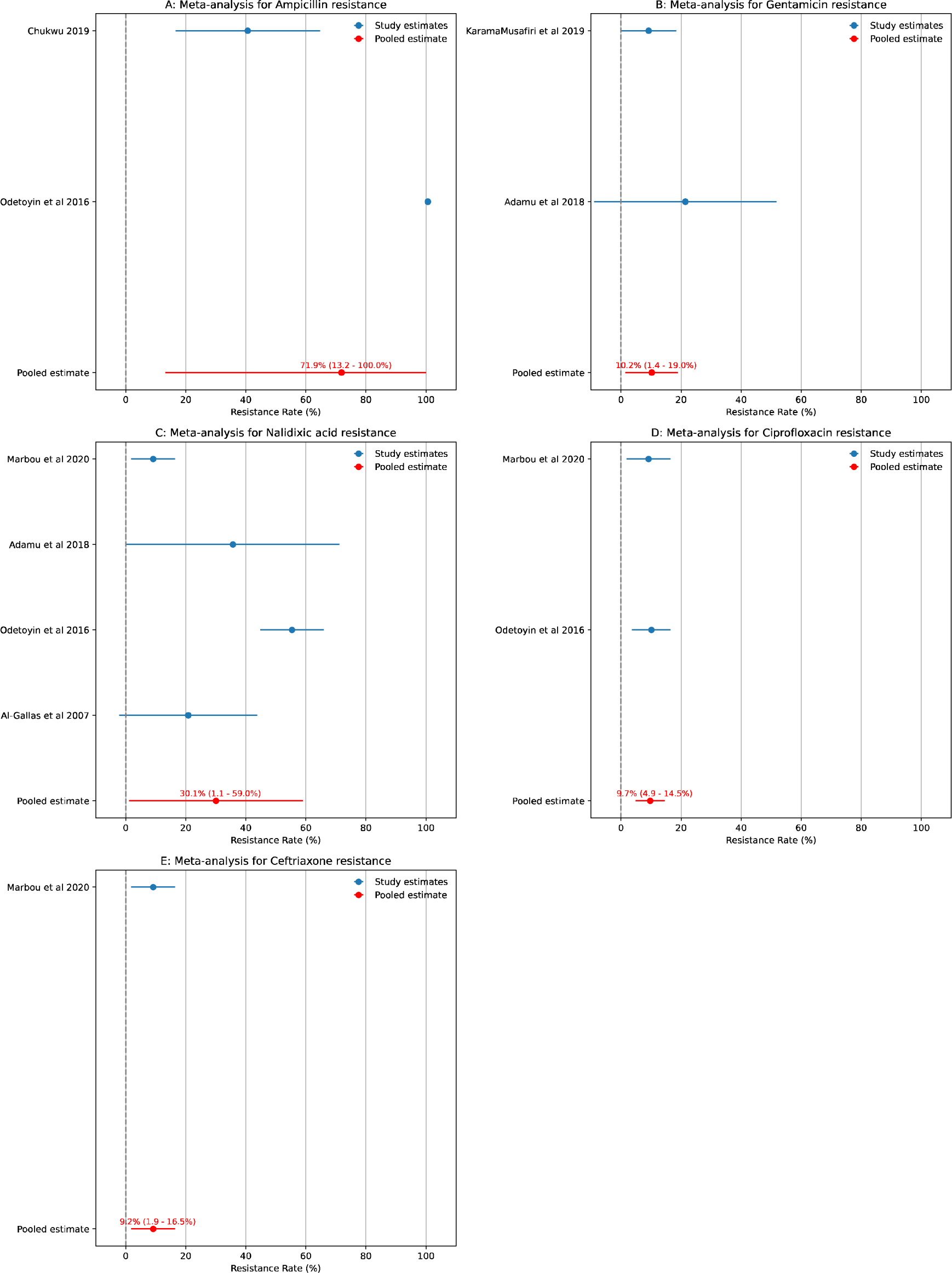
Meta-analysis of antibiotic resistance prevalence in STEC. The figure presents a series of subplots, each corresponding to the prevalence of resistance to a specific antibiotic in STEC samples from various studies. Each blue dot represents the proportion of isolates showing resistance in a particular study. The horizontal position of the dot indicates the percentage of isolates that were resistant, while the dot’s vertical position corresponds to a specific study. Horizontal lines extending from each dot represent the 95% confidence interval of the resistance proportion for that study. The red dot in the first position of each subplot represents the pooled estimate of the resistance rate from a random effects model. The horizontal lines connected to the red dot indicate the 95% CIs of the pooled estimate. Beside each pooled estimate dot is a label specifying the exact resistance percentage and the corresponding 95% CIs.

**Supplementary Figure 2:**
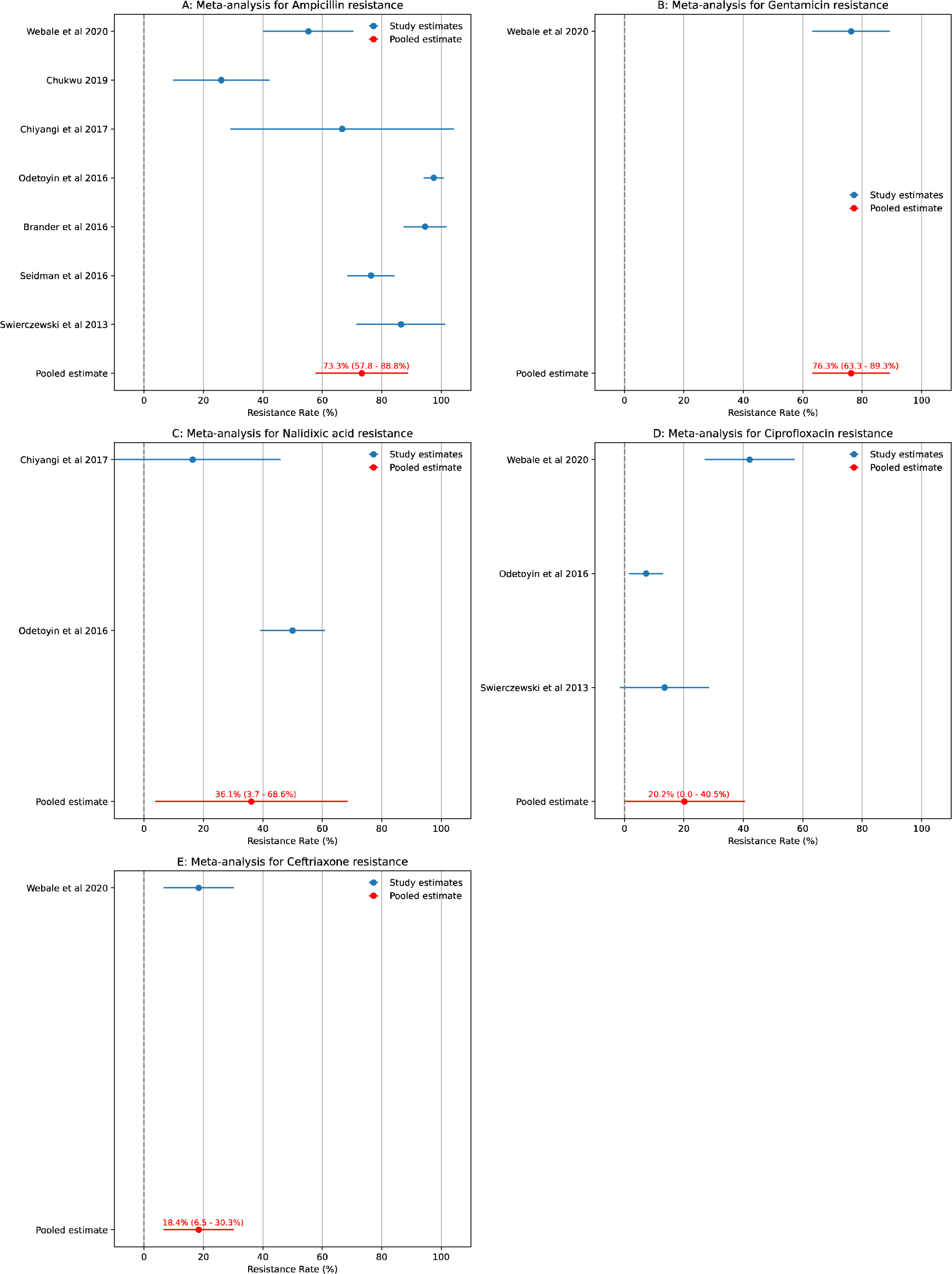
Meta-analysis of antibiotic resistance prevalence in ETEC. The figure presents a series of subplots, each corresponding to the prevalence of resistance to a specific antibiotic in ETEC samples from various studies. Each blue dot represents the proportion of isolates showing resistance in a particular study. The horizontal position of the dot indicates the percentage of resistant isolates, while the dot’s vertical position corresponds to a specific study. Horizontal lines extending from each dot represent the 95% confidence interval of the resistance proportion for that study. The red dot in the first position of each subplot represents the pooled estimate of the resistance rate from a random effects model. The horizontal lines connected to the red dot indicate the 95% CIs of the pooled estimate. Beside each pooled estimate dot is a label specifying the exact resistance percentage and the corresponding 95% CIs.

**Supplementary Figure 3:**
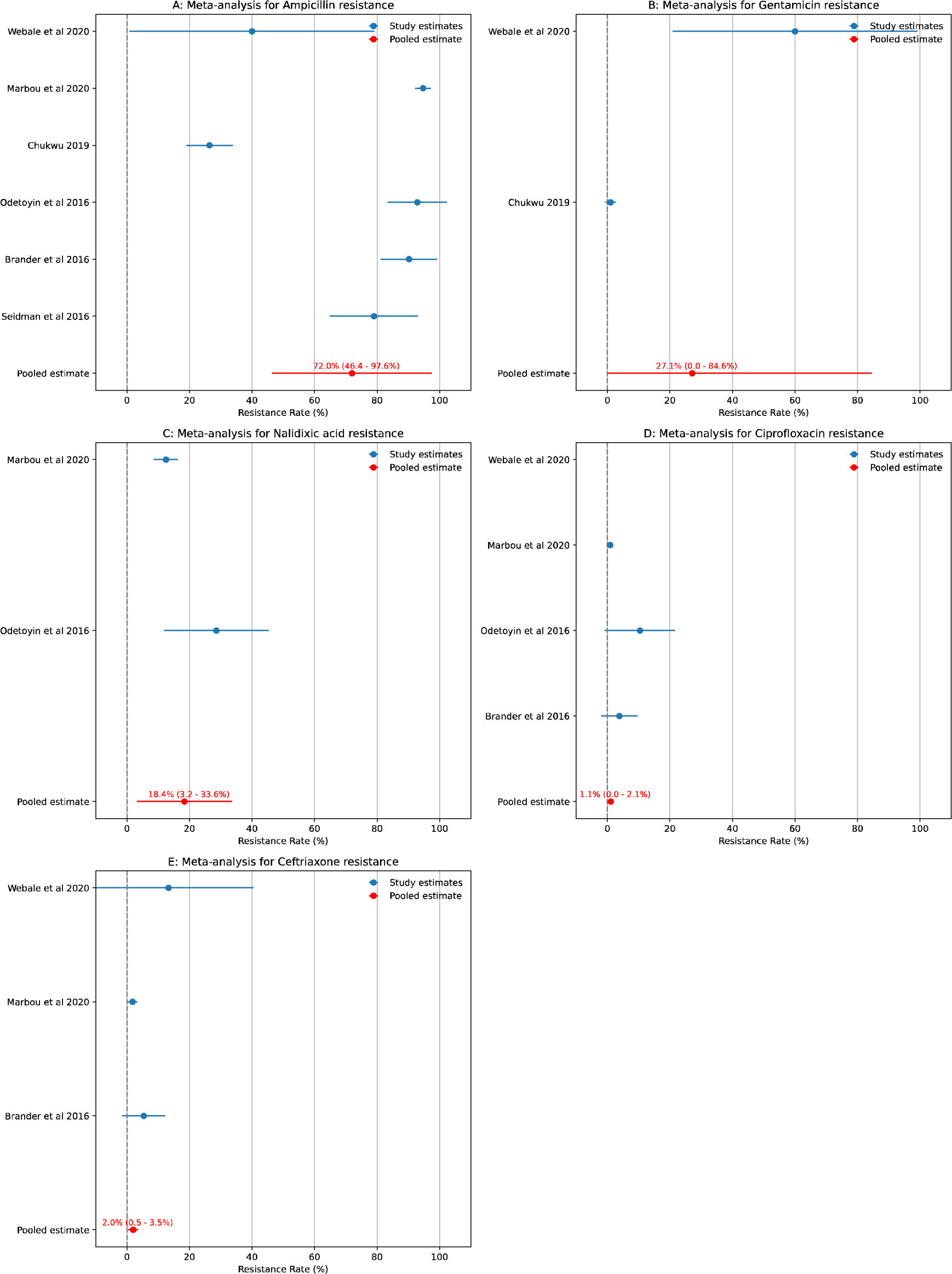
Meta-analysis of antibiotic resistance prevalence in EPEC. The figure presents a series of subplots, each corresponding to the prevalence of resistance to a specific antibiotic in EPEC samples from various studies. Each blue dot represents the proportion of isolates showing resistance in a particular study. The horizontal position of the dot indicates the percentage of resistant isolates, while the dot’s vertical position corresponds to a specific study. Horizontal lines extending from each dot represent the 95% confidence interval of the resistance proportion for that study. The red dot in the first position of each subplot represents the pooled estimate of the resistance rate from a random effects model. The horizontal lines connected to the red dot indicate the 95% CIs of the pooled estimate. Beside each pooled estimate dot is a label specifying the exact resistance percentage and the corresponding 95% CIs.

**Supplementary Figure 4:**
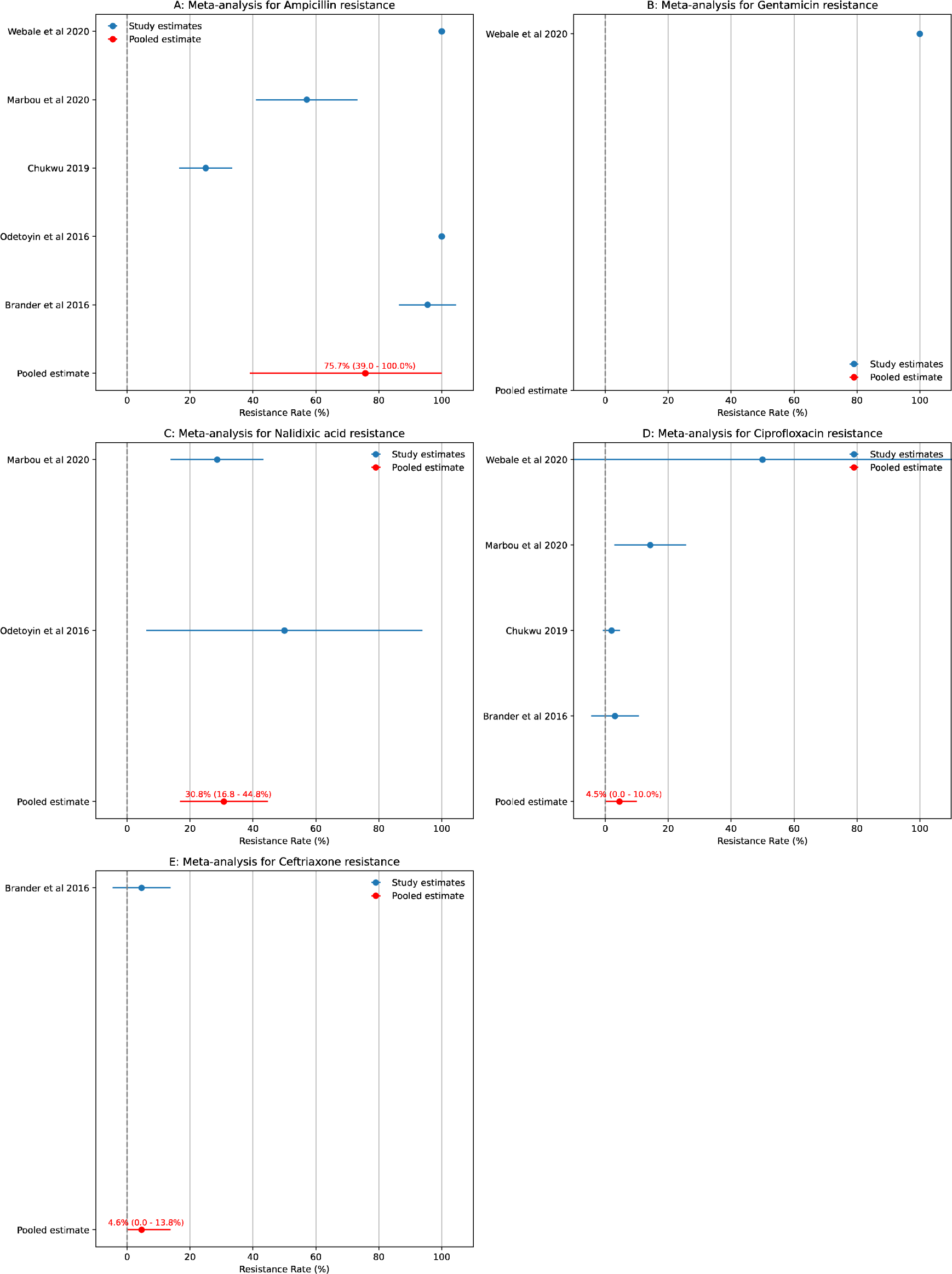
Meta-analysis of antibiotic resistance prevalence in EIEC. The figure presents a series of subplots, each corresponding to the prevalence of resistance to a specific antibiotic in EIEC samples from various studies. Each blue dot represents the proportion of isolates showing resistance in a particular study. The horizontal position of the dot indicates the percentage of resistant samples, while the dot’s vertical position corresponds to a specific study. Horizontal lines extending from each dot represent the 95% confidence interval of the resistance proportion for that study. The red dot in the first position of each subplot represents the pooled estimate of the resistance rate from a random effects model. The horizontal lines connected to the red dot indicate the 95% CIs of the pooled estimate. Beside each pooled estimate dot is a label specifying the exact resistance percentage and the corresponding 95% CIs.

**Supplementary Figure 5:**
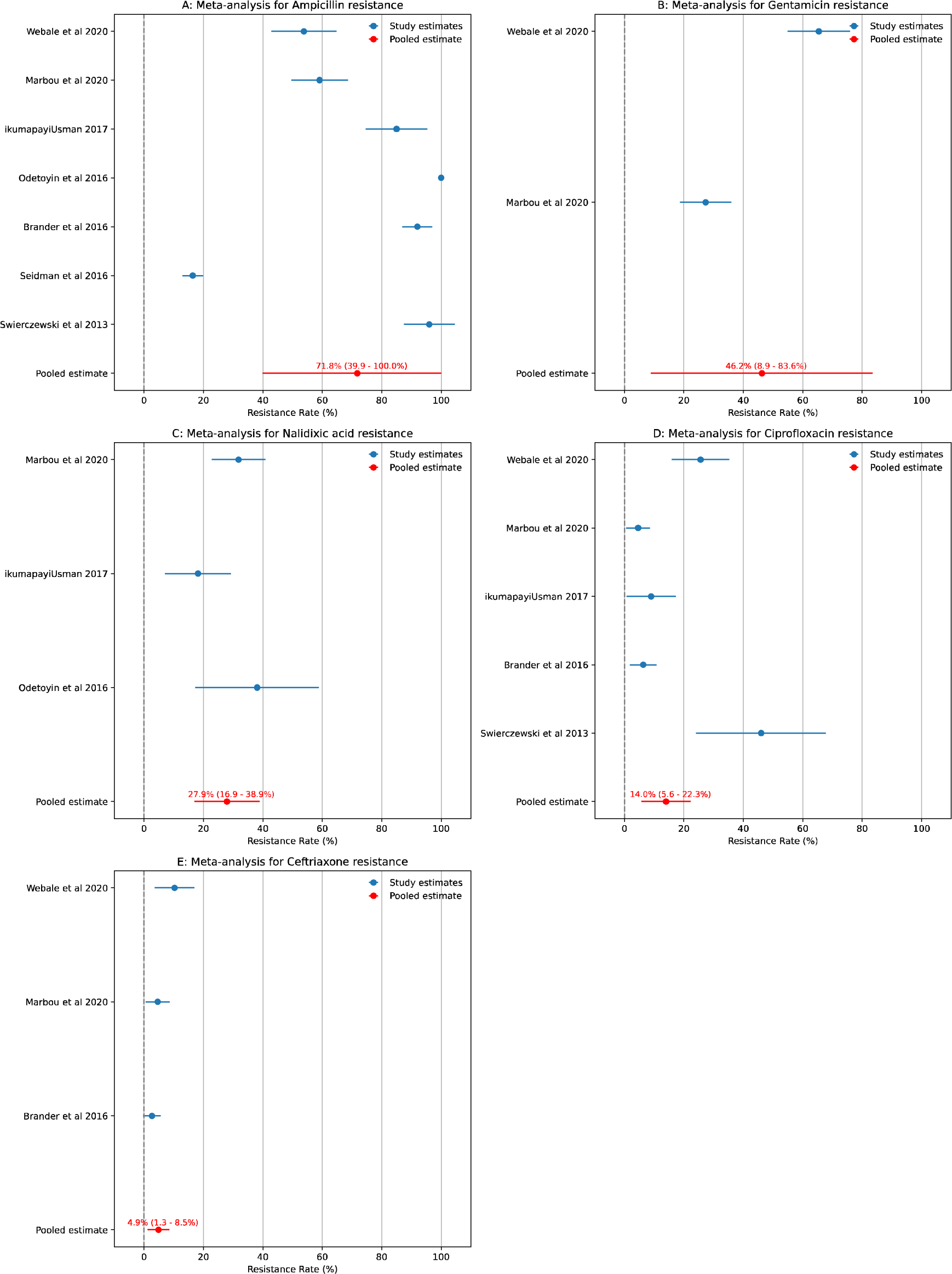
Meta-analysis of antibiotic resistance prevalence in EAEC. The figure presents a series of subplots, each corresponding to the prevalence of resistance to a specific antibiotic in EAEC samples from various studies. Each blue dot represents the proportion of isolates showing resistance in a particular study. The horizontal position of the dot indicates the percentage of samples that were resistant, while the dot’s vertical position corresponds to a specific study. Horizontal lines extending from each dot represent the 95% confidence interval of the resistance proportion for that study. The red dot in the first position of each subplot represents the pooled estimate of the resistance rate from a random effects model. The horizontal lines connected to the red dot indicate the 95% CIs of the pooled estimate. Beside each pooled estimate dot is a label specifying the exact resistance percentage and the corresponding 95% CIs.

